# Morphology-Driven Inference of Patient-Specific Pathophysiological States Enables Precision Treatment in Chronic Spontaneous Urticaria

**DOI:** 10.64898/2026.01.15.26344235

**Authors:** Sungrim Seirin-Lee, Takahiro Hiraga, Hiroshi Ishii, Ryo Saito, Daiki Matsubara, Shunsuke Takahagi, Michihiro Hide

**Affiliations:** Institute for the Advanced Study of Human Biology (ASHBi), Kyoto University Institute for Advanced Study, Kyoto University, Kyoto 606-8315, Japan; Department of Mathematical Medicine, Graduate School of Medicine, Kyoto University, Kyoto 606-8315, Japan; Research Center of Mathematics for Social Creativity, Research Institute for Electronic Science, Hokkaido University, Sapporo 060-0812, Japan; Department of Dermatology, Institute of Biomedical & Health Sciences, Hiroshima University, 1-2-3 Kasumi, Minami-ku, Hiroshima 734-8551, Japan; Department of Dermatology, JA Hiroshima General Hospital, Hiroshima, 738-8503, Japan; Hiroshima City Hospital Organization, Hiroshima 730-8518, Japan; JST CREST, 4-1-8 Honcho, Kawaguchi, Saitama 332-0012, Japan

**Keywords:** Chronic spontaneous urticaria, Skin eruption morphology, Mathematical modeling, Parameter inference, Topological data analysis, Machine learning

## Abstract

Skin diseases manifest as visually observable eruption patterns, making image-based assessment a central component of dermatological diagnosis. While recent artificial intelligence (AI)–based approaches have achieved remarkable progress in classifying skin diseases from images, their utility remains largely limited to pattern recognition tasks, such as disease identification or severity grading. Crucially, most existing AI frameworks operate as black-box classifiers and do not provide interpretable links between eruption morphology and the underlying *in vivo* pathophysiological states, thereby offering limited support for personalized treatment decisions. To date, no practical framework has been established to systematically translate eruption morphology into mechanistic insights or treatment-relevant predictions for inflammatory skin diseases such as chronic urticaria.

Here, we propose a novel integrative framework that infers patient-specific pathophysiological states directly from skin eruption morphology. Our approach unifies mechanistic mathematical modeling with data science that encompasses machine learning and topological data analysis, together with *in vitro* experiments and clinical data into a single coherent system. By constructing a mathematical model that explicitly links disease pathophysiology to eruption morphology, we develop a computational parameter inference tool, the System for Skin Eruption Morphology-based Parameter Inference (SEMPi), that estimates patient-specific physiological parameters directly from real-world skin eruption images. Importantly, these inferred parameters are interpretable in terms of underlying biological processes, enabling direct insight into patient-specific disease states rather than mere image-level classification. Furthermore, by incorporating drug interactions into the mathematical model, our framework enables treatment-response prediction and optimization of individualized therapeutic strategies across multiple drugs. This study introduces a paradigm shift from morphology-based classification toward morphology-driven interpretation of patient physiology, providing a foundation for predictive diagnosis and precision treatment in inflammatory skin diseases.

## 1 Introduction

Chronic spontaneous urticaria (CSU), a major subtype of urticaria, is one of the most intractable human-specific inflammatory skin diseases, characterized by variably shaped wheals and intense pruritus (Zuberbier et al. 2022). The prevalence of chronic urticaria is estimated to be approximately 0.5–1% in the general population, with CSU accounting for the majority of cases and causing substantial impairment in patients’ quality of life over prolonged periods ranging from months to decades. Although CSU pathophysiology has been associated with autoimmune-driven mast cell degranulation, cellular infiltration, activation of the coagulation cascade, and involvement of the complement system (Yanase et al. 2018a,b, 2021), the lack of appropriate animal models and disease-specific biomarkers has severely limited a comprehensive understanding of its *in vivo* mechanisms (He et al. 2026). As a result, therapeutic strategies for CSU remain largely empirical. Indeed, second-generation H_1_ antihistamines, the standard first-line therapy, fail to adequately control symptoms in approximately 30–40% of patients even with dose escalation (Staevska and et al. 2010; Schafer et al. 2025; Zuberbier et al. 2022). While several targeted immunomodulatory agents have recently entered clinical trials, their clinical applicability remains constrained, underscoring the urgent need for approaches that enable reliable prediction of drug efficacy based on patient-specific disease mechanisms (Zuberbier et al. 2024).

Skin diseases, including CSU, manifest as *visible information* in the form of eruption patterns distributed across the body. Accordingly, visual assessment of skin morphology has long served as a cornerstone of dermatological diagnosis. More recently, artificial intelligence (AI)– based image analysis has shown impressive performance in classifying skin diseases and grading disease severity (Biswas et al. 2025). However, most existing AI approaches are fundamentally limited to data-driven pattern recognition. They typically function as black-box classifiers, offering limited interpretability and little insight into the underlying biological processes that give rise to observed morphologies (Daneshjou et al. 2022; Rudin 2019; Topol 2019). Consequently, while such methods may support diagnostic categorization, they do not address a more fundamental clinical challenge: inferring patient-specific *in vivo* pathophysiological states or guiding individualized treatment decisions based on disease mechanisms.

Histological examination of skin biopsy specimens can provide detailed molecular and cellular information, yet this approach is invasive and inherently restricted to static snapshots of tissue states. These limitations have contributed to a long-standing gap in dermatology: the absence of a systematic, non-invasive framework that translates eruption morphology into mechanistic understanding of disease pathophysiology. In inflammatory skin diseases such as CSU, no practical methodology has been established to directly connect detailed eruption geometry with underlying biological networks in a manner that is both interpretable and clinically actionable.

In contrast to purely data-driven approaches, our recent work has demonstrated that eruption morphology in CSU is not merely a superficial manifestation but encodes quantitative information about the underlying pathophysiological state (Seirin-Lee et al. 2020, 2023). By combining mathematical modeling with *in vitro* experiments and clinical data, we previously showed that distinct wheal morphologies correspond to specific dynamical regimes of the CSU pathophysiological network. These findings suggest a conceptual shift: skin eruption patterns can be regarded as macroscopic readouts of patient-specific *in vivo* physiology, rather than as features to be classified in isolation.

Building on this insight, the present study aims to establish a novel methodological framework that infers patient-specific pathophysiological states directly from eruption morphology. We propose an integrative, multi-omics approach that unifies mechanistic mathematical modeling, machine learning, topological data analysis, *in vitro* experiments, and clinical data (Fig.1). First, we construct a mechanistic mathematical model that explicitly links disease pathophysiology to eruption morphology, with model components validated through *in vitro* experiments and overall dynamics assessed by clinical observations (Seirin-Lee et al. 2023). We then develop a computational parameter inference tool that integrates machine learning and topological data analysis to estimate patient-specific model parameters directly from skin eruption images. Importantly, these parameters have explicit physiological interpretations, enabling direct insight into individual disease states rather than opaque image-level predictions.

**Figure 1:**
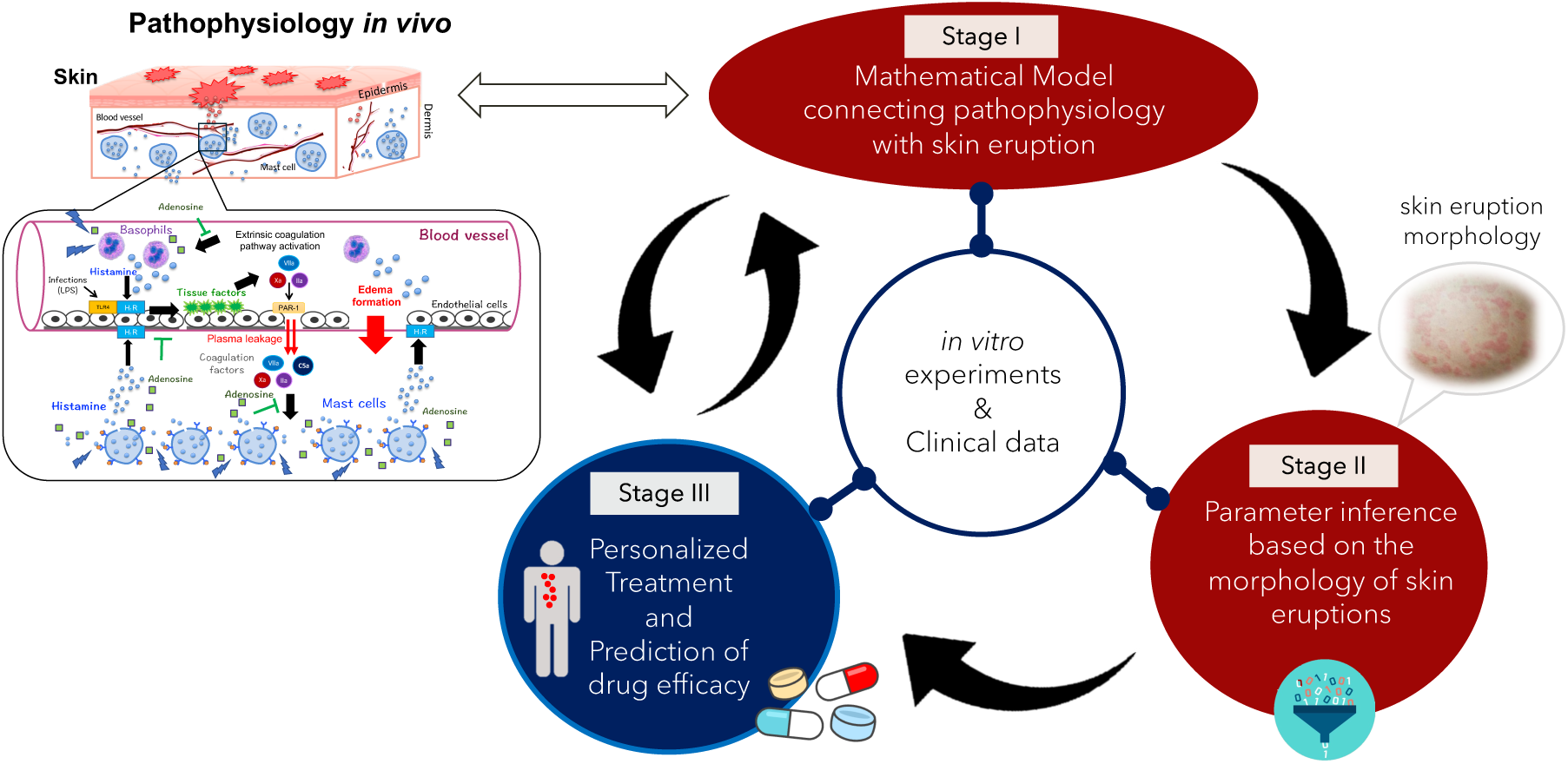
A morphology-driven framework for personalized treatment based on skin eruption patterns. In Stage I, we develop a mathematical model that links disease pathophysiology to skin eruption morphology. In Stage II, we construct a parameter inference system based on the morphological features of skin eruptions. By inferring patient-specific parameter sets from eruption morphology, we subsequently perform *in silico* experiments in Stage III to evaluate drug efficacy and to elucidate how different therapeutic agents act in individual patients.

Finally, we extend the mathematical model to incorporate drug interactions within the pathophysiological network, allowing systematic evaluation of therapeutic responses across multiple agents. By combining patient-specific inferred parameters with a drug-integrated model, our framework enables prediction of drug efficacy and optimization of treatment frequency and dosage on an individual basis. The primary objective of this study is therefore not merely to improve image-based classification, but to establish a generalizable, morphology-driven framework for interpreting patient physiology and supporting personalized therapeutic strategies in inflammatory skin diseases. Together, this work represents a substantive step toward transforming skin eruption morphology from a diagnostic descriptor into a quantitative interface for precision medicine in dermatology.

## 2 Method

### 2.1 Skin eruption segmentation (SEseg) model

To identify the eruption areas of patients with CSU, we carried out a semantic segmentation process using a convolutional encoder-decoder network (e.g., U-Net), which was trained to distinguish skin eruption regions. The network was trained on annotated images reviewed by four dermatologists that served as ground truth, allowing the model to learn characteristic visual features such as color tone, texture irregularity, and boundary morphology of the lesions. See Appendix A for more details.

### 2.2 Persistence homology and persistence image

The persistent diagram (PD) was computed from a filtration constructed by signed distance transform with Euclidian distance (Obayashi et al. 2018). The black/white area in the binary image was shrunk/expanded (negative time direction) and expanded/shrunk (positive time direction). Then, with a filtration of cubical sets of white pixels, we calculated 0th and 1st persistence diagrams using HomCloud (Obayashi 2025). To vectorize the persistence diagrams, we generated the persistence image *ρ* defined by

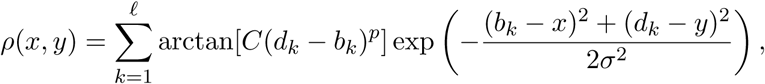

where (*b_k_, d_k_*) is a pair of birth and death values on the persistence diagram, *ℓ* is the total number of points, *C* and *σ* are parameters for avoiding noise data distributed around *y* = *x* on the persistence diagram. The parameters are given to *C* = 1/2, *p* = 1, *σ* = 3.

### 2.3 CSU mathematical model

The CSU mathematical model developed in Seirin-Lee et al. (2023) is based on the pathophysiology of CSU and was validated using *in vitro* experiments and patient clinical data. Therefore, without reiterating the details of the model development, we directly use the model described below.

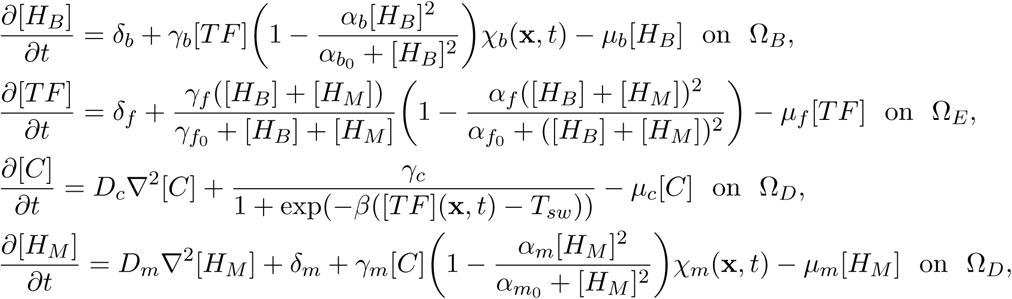

where [*H_B_*](**x**, *t*), [*TF*](**x**, *t*), [*C*](**x**, *t*), and [*H_M_*](**x**, *t*) are concentrations of histamine released from basophils, tissue factor expressed on vascular endothelial cells, activated coagulation factors leaked from blood vessel, and histamine released from mast cells, respectively. Ω*_D_* ⊂ ℝ^2^ is the region of the dermis, Ω*_E_*is the vascular endothelial cells, and Ω*_B_*(⊂ Ω*_E_*) is a random region of the vascular endothelial cells in which basophils affect the endothelial cells. The model is assumed on two dimensional space, Ω*_D_* = Ω*_E_* = [0, *L*] ×[0, *L*] ⊂ ℝ^2^. All parameter values in the model are assumed to be non-negative constants and *T_sw_ >* [*TF*](**x**, 0). *χ_b_*(**x**, *t*) and *χ_m_*(**x**, *t*) are defined as:

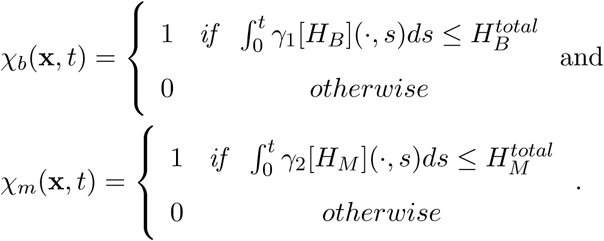

Here, 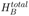 and 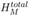 are constants representing the total amounts of histamine contained in basophil and mast cells, respectively. *γ*_1_ and *γ*_2_ are the release rates. The eruption state function on the skin (Ω*_P_* ⊂ ℝ^2^) is given to

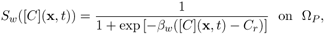

where *C_r_* is a threshold constant value. For detailed descriptions of the parameter meanings and their values, see Table S1 in the Supplementary Information (Seirin-Lee et al. 2023).

### 2.4 Clinical wheal data of CSU patients

Images of skin eruptions (wheals) were collected from 105 patients with CSU who visited the Department of Dermatology at Hiroshima University Hospital between January 1, 2000 and April 15, 2022. The institutional ethics review board approved the study protocol (Ethical Committee for Epidemiology of Hiroshima University, Hiroshima, Japan, approval number: E2020-2388), which involved analyzing images stored in the secured hospital database.

**Figure 2:**
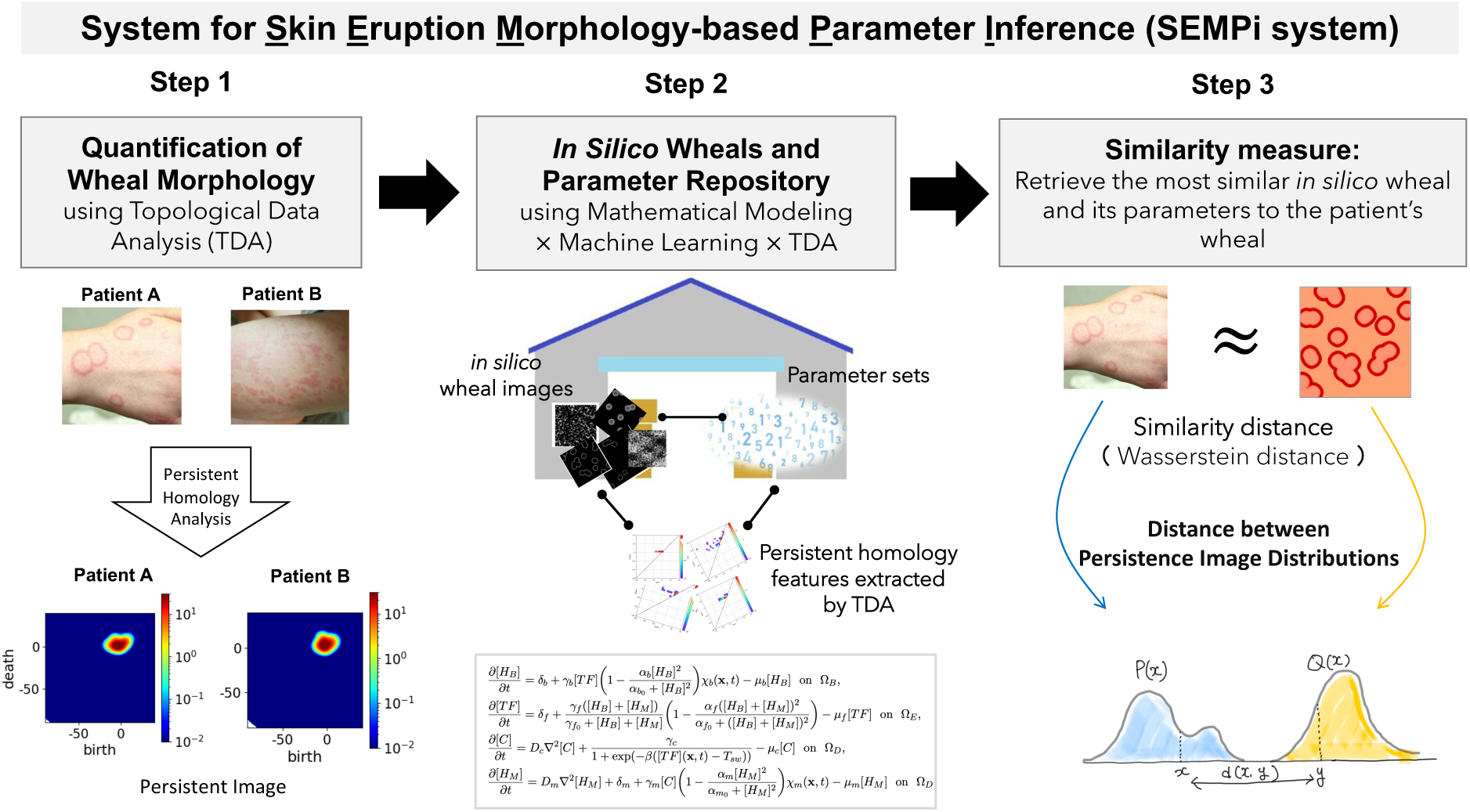
Schematic diagram for skin eruption morphology-based parameter inference (SEMPi) system. SEMPi consists of three steps. Step 1: Skin eruption (wheal) morphology of patients is quantitatively characterized using topological data analysis (TDA) through persistent homology and represented as persistent images. Step 2: A repository of *in silico* wheals and their corresponding parameter sets is generated using the CSU mathematical model by integrating mathematical modeling, machine learning, and TDA. Step 3: The most similar *in silico* wheal to a given patient’s wheal is identified by evaluating the similarity between persistent image distributions. Similarity is quantified using the Wasserstein distance, and the associated parameter set is retrieved as the inferred patient-specific parameter set.

## 3 Result

### 3.1 A system for inferring pathological states based on skin eruption morphology

We first overview the workflow for developing a system for personalized treatment based on skin eruption morphology by inferring parameters representing pathological state of CSU. We call this system Skin Eruption Morphology-based Parameter Inference (SEMPi) system. Since the CSU mathematical model is constructed based on the underlying pathophysiological network of the disease, the model parameters inferred from each patient’s skin eruption (wheal) patterns directly reflect the individual pathophysiological state of CSU.

The system is constructed by the composition of three steps:

**Step 1:** Quantification of skin eruption pattern of patients using topological data analysis, in particular, persistence homology (Otter et al. 2017).

**Step 2:** Create *in silico* data repository by using CSU mathematical model developed by Seirin-Lee et al. (2023).

**Step 3:** Evaluate the similarity of topological data structures between real skin eruption patterns observed in patients and the corresponding *in silico* patterns generated by the CSU mathematical model.

Two key ideas underpin our system. First, to treat eruption patterns within a practical mathematical framework, we transform eruption morphology into quantitative representations. To this end, we employ topological data analysis, specifically persistent homology theory (Turkes et al. 2022). Second, we generate *in silico* eruption patterns using a mathematical model and establish a correspondence between the space of real eruption patterns and the space of *in silico* eruption patterns.

In our previous study (Seirin-Lee et al. 2023), we showed that wheal patterns in CSU can be classified into five distinct types based on eruption geometry criteria (EGe criteria; Fig. 4A). The EGe criteria were developed from feature measures that dermatologists implicitly use to recognize eruption states in clinical settings. Importantly, these feature values suggest that dermatologists do not evaluate precise geometrical shapes of individual eruptions; rather, they capture universal and common morphological characteristics shared across CSU patients. Motivated by this observation, we adopt a topological, rather than a purely geometrical, measure to quantify eruption patterns. Specifically, we employ persistent homology, a core method in topological data analysis that characterizes the shape of data across multiple scales. Persistent homology captures the persistence of topological features, such as connected components, loops, and voids, thereby providing insights into the underlying structure of complex datasets. Here, we omit the methodological details of persistent homology and instead refer the reader to comprehensive reviews for further information (Kumar et al. 2025).

### 3.2 Quantification of skin eruption morphology using topological data analysis

To transform the geometrical feature of skin eruptions of patients to quantitative values, we combined two tools; skin eruption segmentation (SEseg) model based on Convolution Neural Network (CNN) (See Method 2.1 and Appendix A) and the persistent homology of binary image data (Obayashi et al. 2018). Fig. 3A shows the framework of the process.

**Figure 3:**
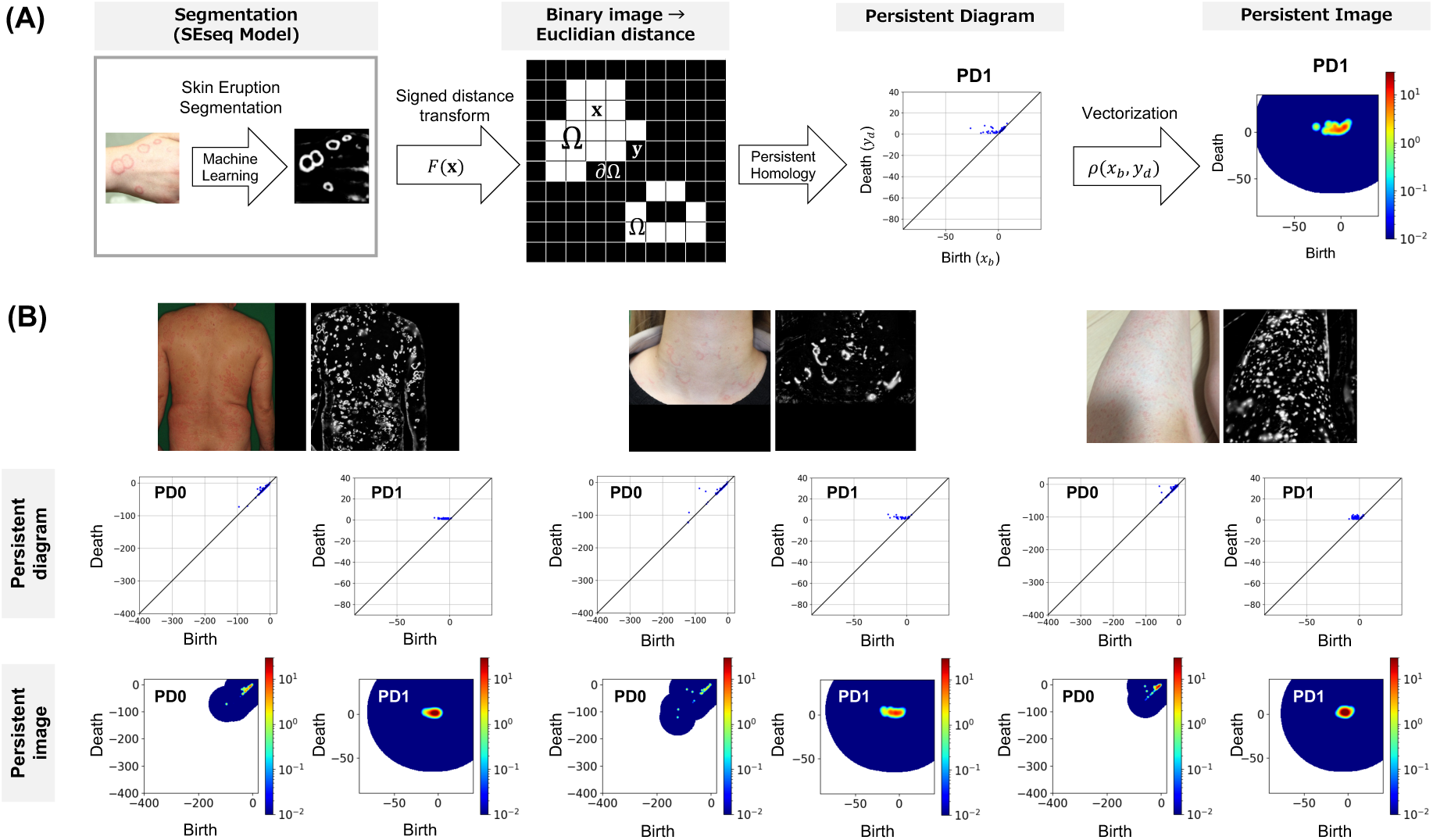
Quantification of skin eruption shapes. (A) Workflow for quantifying skin eruptions using the SEseg model and persistent homology. The signed distance transform *F*(**x**, **y**) is defined as *F*(**x**) = sign(**x**) inf**_y_**_∈*∂*Ω_ ||**x** − **y||**_2_, where Ω denotes the wheal region and *∂*Ω its boundary. The function *ρ*(*x_b_, y_d_*) is defined as described in the Method section 2.2. (B) Representative examples of CSU patients and the corresponding quantification of their skin eruption patterns.

After obtaining grayscale segmentation data of skin eruption regions using our SEseq model, we convert the images into binary format using the Python library imageio.v2. Next, to perform persistent homology analysis, we define a function *F* that assigns an integer value to each pixel in the binary image, corresponding to the signed distance transform with Eculidian distance, as illustrated in Fig.3A. Using a filtration of cubical complexes constructed from the white pixels, we then compute the 0th- and 1st-dimensional persistence diagrams with HomCloud(Obayashi 2025) (see Methods 2.2). Fig. 3B presents representative examples of the quantification of real skin eruption patterns from patients with CSU. Notably, this quantification method does not require specialized imaging equipment; readily obtainable photographs captured with standard smartphones are sufficient for the analysis.

### 3.3 Validation of Morphology Quantification Using Persistent Homology

In this and the following section, we evaluate how effectively the tool based on persistent homology captures the morphological features of skin eruption patterns and assess its reliability as a clinical tool. To this end, we developed a machine learning model based on a fully connected neural network (FCNN) with Rectified Linear Unit (ReLU) activation functions, trained on persistent image representations. We then compared the classification performance of this model with that of six dermatologists using clinical data from CSU patients.

Eruption Geometry (EGe) criteria, previously developed for classifying CSU skin eruption shapes (Seirin-Lee et al. 2023), are illustrated in Fig. 4A. In this framework, CSU skin eruptions are categorized into two primary types, Boundary and Area, then each further divided into five subtypes: *Annular* and *Broken-annular* under the Boundary type, and *Geographic*, *Circular*, and *Dot* under the Area type. In this study, the EGe-based labels assigned through consensus among multiple experienced dermatologists were used as the gold standard for classification. Fig. 4B presents the classification reliability of six dermatologists for 105 patient cases relative to this gold standard. While the classification reliability for the Boundary and Area categories exceeded 80%, the reliability for the five subtypes was below 70%. Moreover, the observed standard deviations indicate substantial inter-observer variability, reflecting the dependence of subtype classification on subjective judgment. These results highlight the intrinsic difficulty of precisely characterizing wheal morphology, even for trained dermatologists, and underscore the need for an objective, mathematically defined measure for more reliable classification.

**Figure 4:**
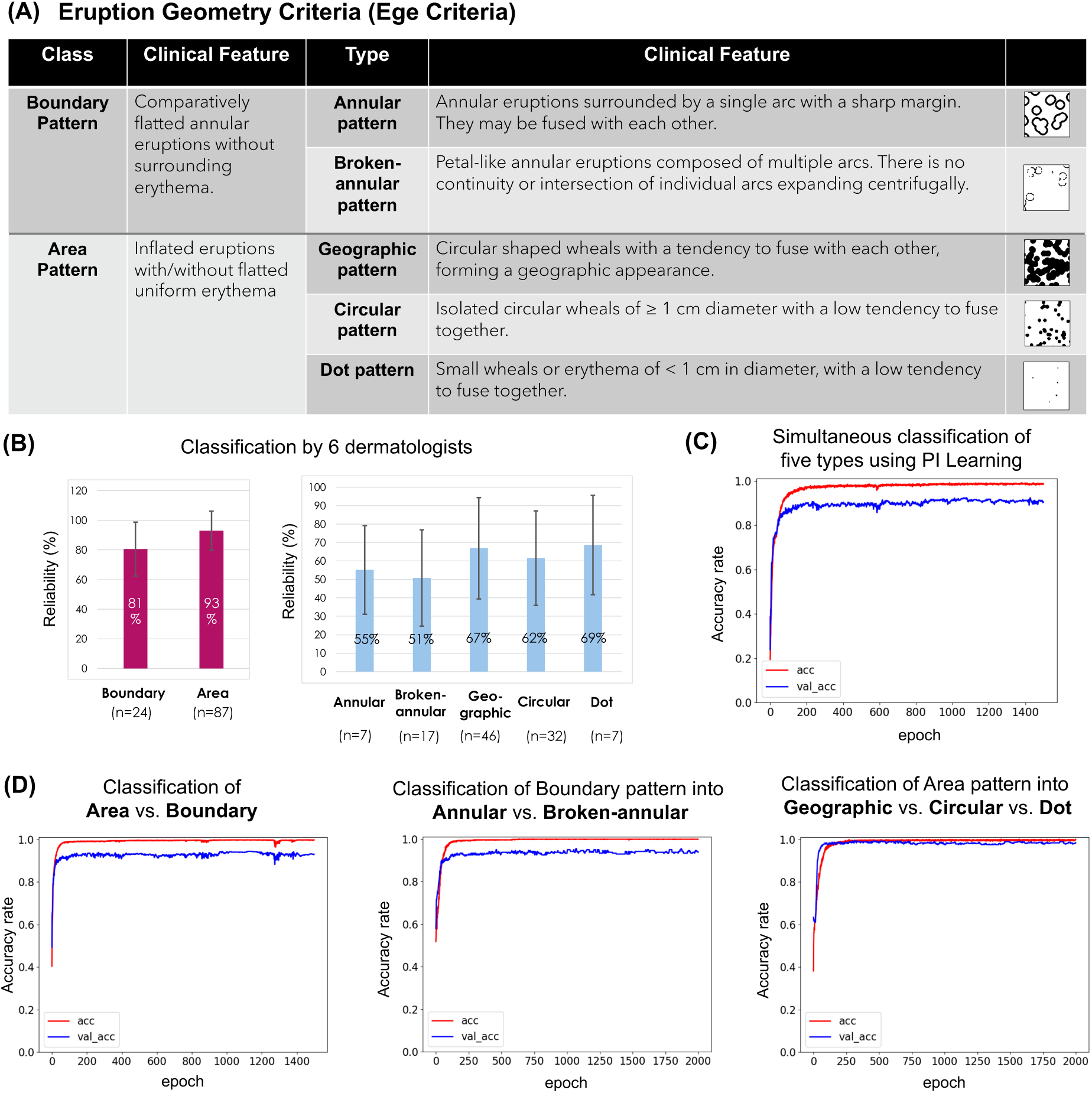
Validation of morphology quantification and classification using persistent homology. (A) Eruption Geometry Criteria for classifying the wheal types of CSU (Seirin-Lee et al. 2023). (B) Classification results by six dermatologists. (C-D)Persistent images of both the 0th and 1st persistent diagrams were used in the machine learning models. The red and blue lines indicate accuracy(acc) and validation accuracy(val acc) rates, respectively. (C) Classification results using a multilayer perceptron machine learning model based on persistent images, simultaneously classifying five wheal types. Mean(acc, val acc)=(0.9834, 0.9050) (D) Classification results using a multilayer perceptron machine learning model based on persistent images. The initial step classifies wheals into two categories: boundary or area (left panel), followed by classification into respective subtypes (middle and right panels). Mean(acc, val acc)=(0.9952, 0.9298), (0.9986, 0.9383), (0.9956, 0.9825) in left, middle, and right panels.

We next analyzed the same 105 CSU patient cases using the workflow illustrated in Fig. 3A. A multilayer perceptron model was trained in a supervised manner using persistent images, with the dermatologist-consensus EGe labels serving as the gold standard. A total of 3,000 training samples were generated from 31 patients by extracting multiple local segments from each patient’s full skin image. Two classification strategies were evaluated: (i) a single-step approach that simultaneously classifies all five subtypes, and (ii) a hierarchical approach that first distinguishes between Boundary and Area types and subsequently classifies the corresponding subtypes. As shown in Fig. 4C, the single-step classification achieved approximately 98% accuracy. The hierarchical approach further improved performance, yielding approximately 99% accuracy for both the Boundary-versus-Area classification (left panel) and the subsequent subtype classification (middle and right panels). These results demonstrate that the machine learning model leveraging persistent homology can reproduce, and in fact exceed, the consistency of human expert classification, by quantitatively capturing the essential morphological features of wheal patterns.

### 3.4 Interpretation of Morphological Features Captured by Persistent Homology in Comparison with Clinical Measures

What features does persistent homology capture to classify wheal types? One of the key motivations for adopting persistent homology in this study, rather than relying solely on conventional AI-based image classification, is its interpretability. While many deep learning approaches achieve high classification accuracy, they often operate as black boxes, providing limited insight into which morphological features drive the classification. In contrast, the persistent homology– based representation allows direct interrogation of the extracted features. By combining dimensionality reduction with inverse analysis, we can explicitly interpret which morphological characteristics are used for classification and examine whether these features are consistent with clinical criteria (EGe Criteria) used by dermatologists.

To this end, we applied Principal Component Analysis (PCA) to the persistent image data to extract key features and performed inverse analysis to interpret the dominant principal components. Specifically, PCA was used to identify the components that contribute most strongly to the clustering of wheal patterns in the persistent image space, and inverse analysis was then conducted to trace these components back to the original patient images.

We first found that the three principal components derived from PCA effectively distinguish between annular and broken-annular patterns (Fig. 5A, left panel). To interpret the morphological features captured by each component, we performed inverse analysis on the top three principal components of the PD0 and PD1 persistent image data (Fig. 5A, right panel). For the PD0 data, the components primarily reflected the non-pattern space, capturing structural features such as the presence or absence of isolated regions (e.g., white circular areas). In contrast, for the PD1 data, the components directly represented morphological characteristics specific to annular and broken-annular patterns (Fig. 5A, right panel, green dotted circles).

**Figure 5:**
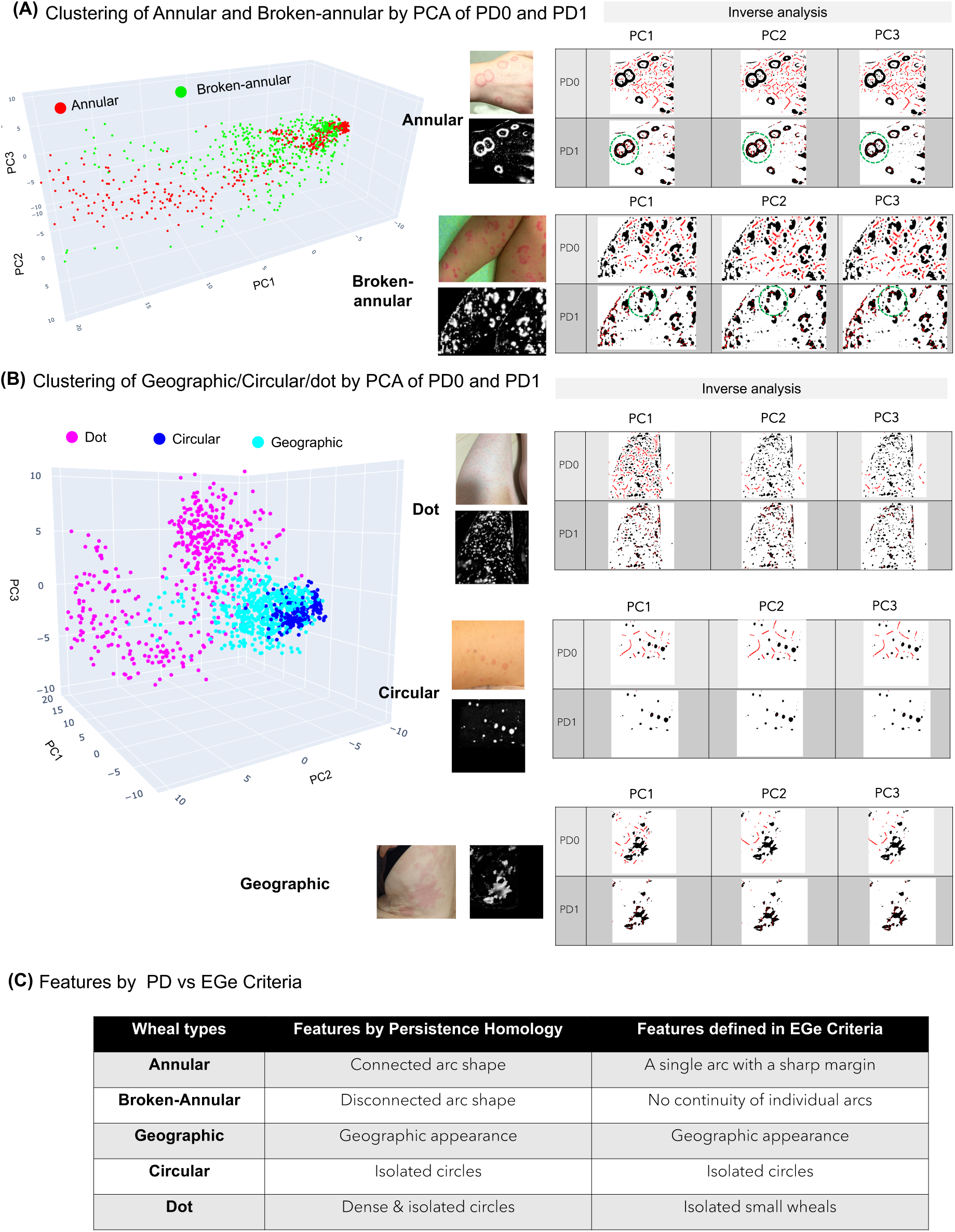
Principle component analysis (PCA) based on persistent image (PI) data and inverse analysis. (A) PCA analysis for annular and broken-annular patient data, along with inverse analysis. (B) PCA analysis for geographic, circular, and dot patient data, along with inverse analysis. (C) Summary of the features based on PCA analysis of PI and EGe Criteria.

We next conducted a similar PCA-based analysis for the geographic, circular, and dot patterns observed in CSU patients (Fig. 5B, left panel). These three patterns were also well separated along the three principal components. Inverse analysis revealed that, in the PD0 data, the primary component (PC1) captured pattern density, enabling clear discrimination of dot patterns from the other two types (Fig. 5B, right panel, PC1 and PD0). Conversely, in the PD1 data, PC1 captured shape-related features that differentiated circular patterns from irregular (geographic) patterns (Fig. 5B, right panel, PC1 and PD1, green dotted circles).

Finally, we summarized the morphological features identified by persistent homology alongside those recognized by dermatologists in Fig. 5C. Notably, the features extracted through persistent homology closely correspond to the EGe Criteria used in clinical classification (Fig. 5D). This correspondence demonstrates that the topological features driving the model-based classification are not arbitrary mathematical constructs, but rather reflect clinically meaningful morphological characteristics implicitly used by experienced dermatologists.

Together, these results highlight a major advantage of the persistent homology–based approach: it not only achieves accurate classification, but also provides an interpretable framework in which the underlying morphological features can be explicitly examined and validated by established clinical criteria. This interpretability distinguishes our approach from black-box AI models and supports its potential utility as a reliable and clinically meaningful tool for objective assessment of CSU eruption morphology.

### 3.5 Morphology-Based Similarity Mapping and Construction of the *in silico* **Wheal Repository**

Here, we introduce a framework that links real wheals observed in CSU patients with *in silico*-generated wheals, enabling the inference of patient-specific parameter sets underlying disease pathophysiology. To formalize this concept, we define a similarity map, 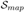, as illustrated in Fig. 6A, 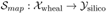, such that

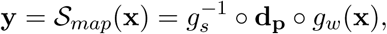

where 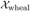 and 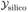 denote the parameter spaces corresponding to real patient wheals and *in silico* wheals, respectively. The mappings *g_w_* and *g_s_* are injective functions that transform a wheal image into its corresponding persistent image representation in the patient and *in silico* spaces. The function **d_p_** maps the persistent image space of patient wheals 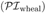 to that of *in silico* wheals 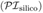, such that similarity between the two is preserved according to a predefined metric, which is specified below.

**Figure 6:**
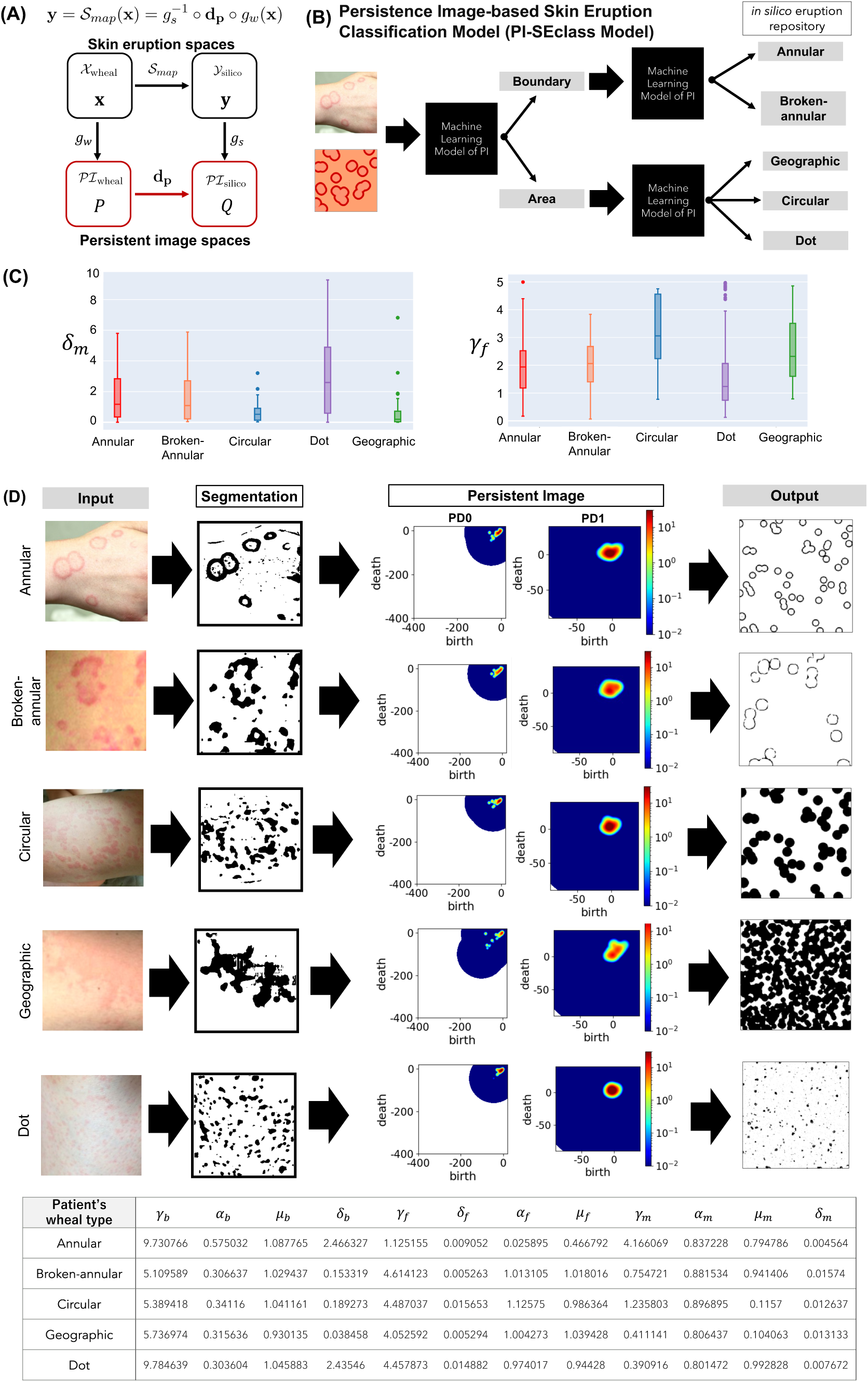
Creation of *in silico data* repository and similarity measure. (A) Definition of similarity map for inferring patient-specific parameter set based on wheal shapes. (B) PI-based Skin Eruption Classification Model(PI-SEclass Model). The process of a machine learning model of persistent images to create each repository of each type of wheals. With this classification process, a patient’s wheal image selects one wheal type of repository and then picks up the parameter values using the similarity measure (**d***_p_*). (C) Examples of the parameter regions of *δ_m_* and *γ_f_* for local sampling derived from global sampling. (D) The representative examples of parameter inference for CSU patients using the SEMPi system.

To improve inference accuracy, we define a pattern-specific similarity map 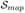 for each wheal type. For this purpose, we first construct a classification model based on persistent image data, as shown in Fig.6B. Two classifiers are trained separately: one using patient-derived data and the other using *in silico* data generated from the mathematical model. We refer to these classifiers collectively as the Persistence Image–based Skin Eruption Classification model (PI-SEclass; see AppendixB for details).

Next, we generate an *in silico* wheal space 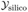 while explicitly preserving the correspondence among three components: *in silico* wheal images, their underlying parameter sets, and the associated persistent image representations 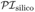. This construction yields an *in silico* repository that enables direct comparison between real and simulated wheals in both morphological and parameter spaces.

To obtain a sufficiently rich and biologically plausible *in silico* dataset, we adopted a two-stage sampling strategy consisting of global and local sampling. In the global sampling stage, all free parameters in the mathematical model were sampled within a predefined range,

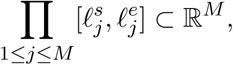

where *M* denotes the total number of parameters. Parameter sets were randomly sampled using a uniform distribution based on the eFAST method (Saltelli et al. 1999). The resulting *in silico* wheals were classified into five wheal types using the PI-SEclass model. Subsequent box-plot analyses allowed us to identify parameter ranges characteristic of each wheal type (Fig. 6C). For each parameter, the minimum and maximum values defining these ranges were denoted as 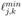 and 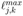, where *k* ∈ [1, 5] indexes the wheal types (see Fig.S3 in AppendixC). Notably, boundary-type patterns (Annular and Broken-annular) and area-type patterns (Circular and Geographic) were distinguished by the parameter *δ_m_*, while the three area-type patterns (Circular, Dot, and Geographic) were further differentiated by *γ_f_*. These observations indicate that each wheal type is characterized by distinct and relatively narrow parameter ranges.

Based on these results, we subsequently performed local sampling by restricting parameter values to the type-specific ranges identified during global sampling. Higher sampling densities were assigned to key parameters, allowing finer resolution within their respective ranges. This approach yielded additional *in silico* samples with more precisely resolved parameter values, improving the accuracy of the similarity mapping. Note that, although wheal formation is inherently a spatiotemporal process, we observed that temporal evolution primarily affects wheal size rather than its fundamental morphological pattern. While changes in wheal size alter topological features captured by persistent homology (see Appendix D and Fig. S2), the overall pattern structure remains preserved. Accordingly, we generated *in silico* wheals at multiple fixed time points using identical parameter sets to account for temporal variability in the analysis.

### 3.6 Topological Similarity Measure for Patient-Specific Parameter Inference

We finally define a quantitative similarity measure between real and *in silico* wheals using the Wasserstein distance between persistent image representations. Specifically, the *q*-Wasserstein distance is defined as

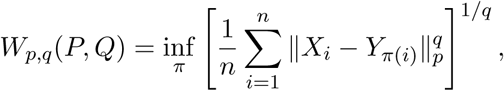

where *P* and *Q* are the distributions of persistent images of wheals from CSU patients and *in silico* wheals, respectively. Here, *π* represents all permutations of *n* elements, || ·||*_p_* denotes the *L_p_* norm, and *q*(≥ 1) specifies the order of the Wasserstein distance. In this study, we primarily adopt *q* = 1. The value of *n* is determined by the resolution of the persistent images.

To eliminate scale-dependent variations in persistent image data, we further define a normalized similarity measure as

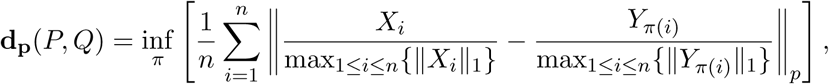

For a given patient wheal pattern *P*, we identify the most similar *in silico* wheal *Q* from the repository by minimizing the combined distances of the zeroth and first persistent homology components (PD0 and PD1):

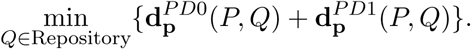

This formulation allows us to simultaneously incorporate both connected-component information (PD0) and loop-related morphological features (PD1) of skin eruptions.

Representative examples of patient’s *in silico* wheal matching obtained using the current repository are shown in Fig. 6D. Importantly, the *in silico* repository can be expanded without limitation by introducing finer parameter discretization within defined parameter ranges and by incorporating additional temporal snapshots. Such expansion is expected to further enhance the precision of patient-specific parameter inference for CSU.

### 3.7 Application of SEMPi to Personalized Treatment Prediction

Here, we demonstrate a proof-of-concept application of our framework by evaluating the effects of an *H*_1_ antihistamine in two representative CSU patients inferred in Fig.6D, focusing on the Annular and Circular wheal types. *H*_1_ antihistamines exert their therapeutic effect by inhibiting histamine binding to the histamine *H*_1_ receptor (*H*_1_R) on the membranes of endothelial cells in blood vessels (Fig.7A).

**Figure 7:**
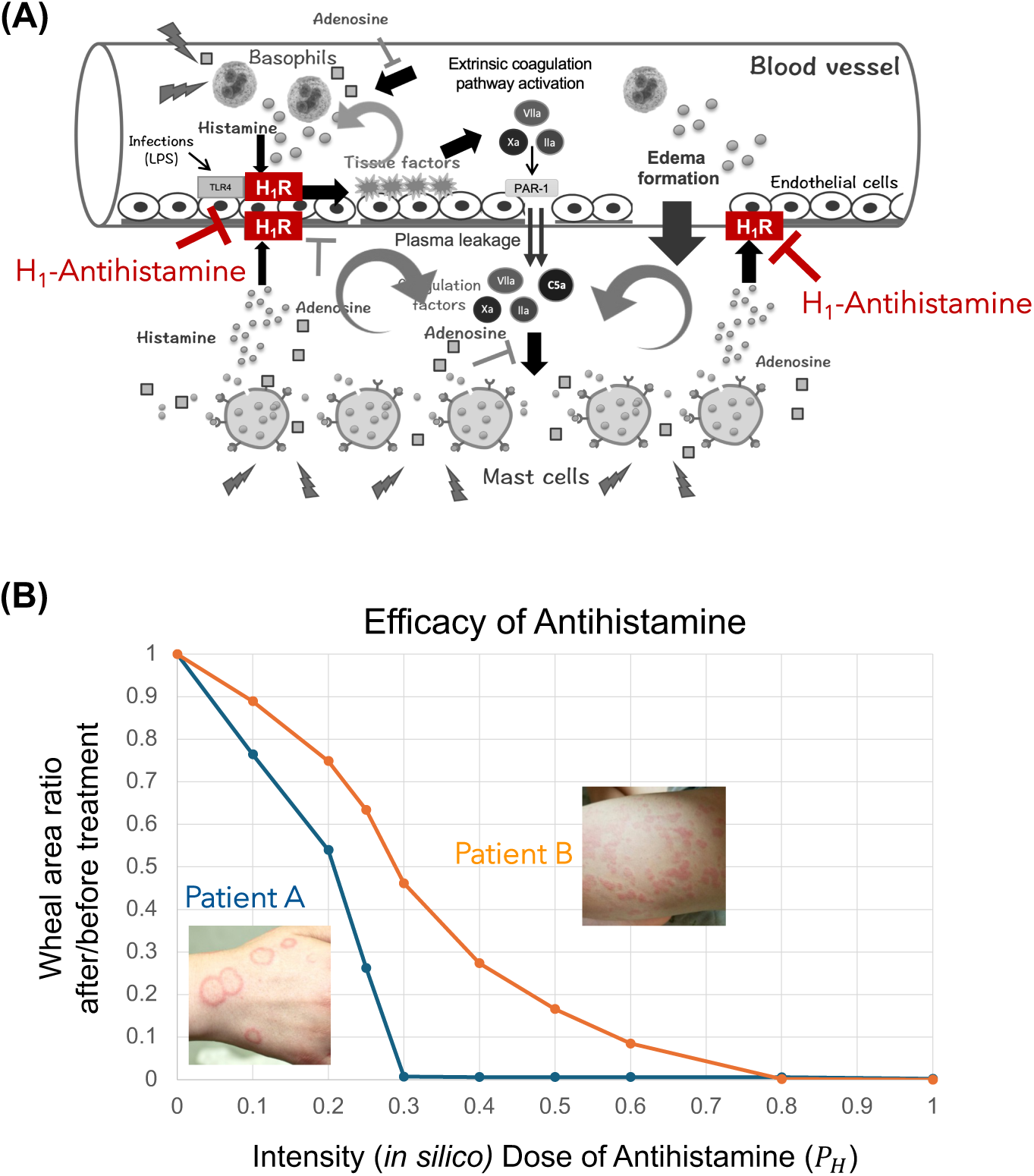
*in silico* Evaluation of antihistamine efficacy using SEMPi system. The parameter values are provided as estimated from the annular (Patient A) and circular (Patient B) types, respectively, as indicated in Fig. 6D. Patient B shows a weaker response to increasing doses of antihistamine compared with Patient A.

To model this mechanism, we extended the CSU mathematical model to incorporate the pharmacological action of *H*_1_ antihistamines as follows:

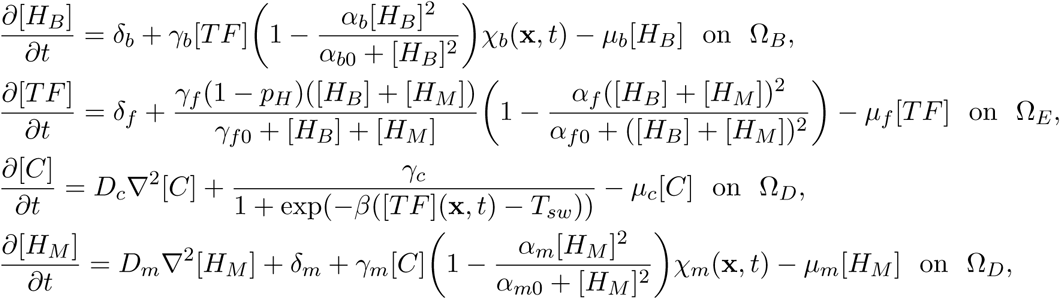

where *p_H_* represents the *in silico* intensity (dose) of the *H*_1_ antihistamine and takes values in the range [0, 1]. Specifically, *p_H_* = 0 corresponds to the pretreatment condition, whereas *p_H_* = 1 represents complete inhibition of *H*_1_R-mediated signaling. To quantitatively assess treatment efficacy, we define the following metric:

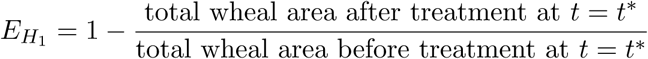

The simulation results are shown in Fig. 7B. Notably, the two patients exhibited markedly different responses to antihistamine treatment. For Patient A, sufficient therapeutic efficacy was achieved with a relatively low dose of antihistamine, whereas Patient B required a substantially higher dose to approach complete remission.

In this analysis, we examined an *H*_1_ antihistamine that is currently used as a standard first-line therapy in clinical practice. Despite this simplicity, the results clearly demonstrate pronounced inter-patient variability in drug response, underscoring the limitations of uniform treatment strategies and the importance of personalized therapeutic decision-making in CSU. Importantly, because our framework is based on a mechanistic mathematical model, it can be readily extended to incorporate multiple therapeutic agents targeting different components of the pathophysiological network. This capability opens the possibility of systematically predicting and optimizing patient-specific responses to diverse treatment options, rather than being restricted to a single drug.

## 4 Discussion

Despite its high prevalence and substantial clinical burden, the management of chronic urticaria remains challenging. Although second-generation antihistamines are recommended as first-line therapy, approximately 30–40% of patients remain unresponsive even after dose escalation (Schafer et al. 2025; Zuberbier et al. 2022). For such patients, international guidelines advocate a stepwise escalation strategy, including second-line therapies with omalizumab, followed by third-line treatment with ciclosporin ciclosporin (Zuberbier et al. 2022). Moreover, several emerging therapies such as biologics (e.g., dupilumab) and small-molecule inhibitors (e.g., remibrutinib) are considered as therapeutic options (Zuberbier et al. 2024). However, the widespread clinical use of these therapies is often limited by high costs, safety concerns, and marked inter-individual variability in therapeutic response (Kolkhir et al. 2022). Indeed, accumulating evidence indicates that differences in autoimmune and immunological profiles among CSU patients are associated with heterogeneous treatment outcomes, including limited efficacy of omalizumab in certain subgroups and comparatively favorable responses to ciclosporin in others (Lang et al. 2025; Lee et al. 2024; Ridge et al. 2025). Importantly, no currently available therapy achieves complete disease control in all patients.

Together, these observations underscore the substantial heterogeneity in the underlying pathophysiological states of CSU and highlight a fundamental limitation of current clinical practice: treatment selection remains largely empirical and insufficiently informed by patient-specific disease mechanisms. From this perspective, the variable and incomplete efficacy of existing therapies emphasizes the urgent need for approaches that can stratify patients according to their underlying pathophysiology and thereby enable more rational, personalized treatment strategies (Zuberbier et al. 2024).

To address this unmet need, we developed an innovative parameter inference system based on the morphological features of patients’ wheals (Fig. 2, SEMPi system). This system infers the *in vivo* pathological state of individual patients using only wheal images acquired with widely accessible devices, such as personal smartphones. Importantly, this approach imposes no physical burden on patients, unlike invasive procedures such as skin biopsies, and substantially reduces the economic costs associated with repeated clinical and pathological testing. Moreover, by extending the underlying mathematical model to incorporate pathological mechanisms targeted by emerging therapeutics, the SEMPi framework enables rapid *in silico* evaluation of drug efficacy using patient-specific parameters. While this study focuses on CSU, the methodology is potentially applicable to other inflammatory skin diseases characterized by spatially organized eruption patterns, provided that a mathematical model linking disease pathophysiology to eruption morphology is available.

A key conceptual foundation of our approach lies in the biological mechanism underlying CSU eruption formation. The onset of CSU wheals is hypothesized to be driven by a basophil-mediated positive feedback loop, initiated by random adhesion of basophils to endothelial cells within capillaries, followed by histamine release (Seirin-Lee et al. 2025). Because the specific locations of wheals are determined by this stochastic initiation process, similarity among eruption patterns should be defined not by pixel-level correspondence or data-assimilation-based similarity, but by universal morphological characteristics shared within each wheal type. This notion is supported by our finding that features extracted using persistent homology closely align with the EGe criteria employed by dermatologists for clinical classification (Seirin-Lee et al. 2023) (Fig. 5). By adopting this morphology-based perspective, we established a direct correspondence between the parameter space of real wheal shapes observed in CSU patients and that of *in silico* wheals generated by our mathematical model. Because the inferred model parameters encode patient-specific pathophysiological states, this framework enables not only the prediction and evaluation of treatment efficacy but also a deeper mechanistic understanding of the origins of patient-specific symptoms. This interpretability represents a critical advantage over conventional AI-based approaches, which typically focus on morphological classification without direct linkage to underlying biological mechanisms (Daneshjou et al. 2022; Rudin 2019; Topol 2019).

In addition, this study introduces an objective mathematical framework for evaluating the similarity of eruption patterns. To date, most theoretical studies on pattern formation have assessed model validity by visually comparing *in silico* patterns with real-world observations. Such comparisons, however, are inherently subjective and lack reproducibility. Importantly, similarity of eruption patterns is conceptually distinct from geometric similarity in terms of precise shape or size; rather, it reflects abstract or conceptual resemblance that captures essential structural features. In this context, we applied topological data analysis, specifically persistent homology, which proved highly effective in quantifying the characteristic morphological features of urticaria eruptions. Persistent homology is an algebraic method that captures topological information such as connectivity and holes, and has been widely applied to analyze data structure in diverse fields ranging from genetics and materials science, to medicine (Amezquita et al. 2020; Kumar et al. 2025; Hernandez-Lemus 2025; Skaf and Laubenbacher 2022). However, its application to defining pattern similarity itself has been limited. Our results demonstrate that persistent homology provides a principled and clinically meaningful measure of eruption morphology, closely mirroring how dermatologists assess wheal patterns. Establishing such objective measures of pattern similarity represents an important step toward bridging the gap between theoretical models of pattern formation and their practical application in life sciences.

Several limitations and future directions should be noted. Improving the accuracy of parameter estimation will require the incorporation of larger and more diverse clinical datasets, as well as further expansion of the *in silico* repository. At the current stage, we have developed an initial version of the parameter inference tool, which can be systematically refined through denser parameter sampling and the inclusion of additional temporal snapshots. Furthermore, collecting wheal images from CSU patients at multiple time points is expected to enhance both the performance of the machine learning components and the robustness of the parameter inference process.

In conclusion, no general framework in dermatology has previously linked the morphological features of skin eruptions in inflammatory skin diseases directly to precision treatment strategies. Building on our earlier work demonstrating the association between CSU eruption morphology and underlying pathophysiological mechanisms (Seirin-Lee et al. 2023), this study substantially advances the concept of morphology-based disease stratification by introducing a transformative framework that infers patient-specific pathophysiological states directly from eruption geometry. By converting visible skin manifestations into quantitative, mechanistic descriptors of disease states, our approach bridges a critical gap between clinical observation and precision therapeutics. Beyond CSU, this framework establishes a general paradigm for integrating mathematical modeling, data-driven inference, and clinical imaging, thereby opening new avenues for personalized treatment strategies across inflammatory skin diseases. More broadly, this work highlights the power of interdisciplinary methodologies in reshaping how dermatological diseases are understood, diagnosed, and treated.

## Data Availability

All data produced in the present study are available upon reasonable request to the authors

## Data availability

All relevant data are included in the manuscript and the supporting information files.

## Code availability

Numerical code is made available online on GitHub (https://github.com/seirin-lee/urticaria-code) and all other data are available from the corresponding author (SSL) on reasonable request.

## Pantent

Some of the techniques presented in this study are currently under patent application (Application No. 2025-111616).

## Author Contributions

SSL initiated, designed, conducted, and supervised the research. SSL developed all mathematical models used in this study. SSL, TH, and HI analyzed the data. TH developed the machine learning and segmentation models. HI initially developed the Python code using HomeCloud, and TH further analyzed the data. SSL developed the numerical algorithms for the *in silico* repository data and codes for mathematical models. DM, RS, ST, and MH collected and provided the clinical data. SSL drafted the initial version of the manuscript and was primarily responsible for major revisions. All authors reviewed and revised the final version.

## Acknowledgements

This study was supported by the Japan Science and Technology Agency(JST) CREST (JP-MJCR2111) and Grant-in-Aid for Transformative Research Areas(A) (22H05110) to SSL.

## Appendix

### A Skin eruption segmentation (SEseg) model

In segmentation tasks, we constructed and trained an encoder-decoder network model using VGG16 (Simonyan and Zisserman 2015) as the encoder, which offers good performance while remaining computationally feasible. Both the input and output of the learning model are given to images. Representative examples include SegNet (Badrinarayanan et al. 2016), Fully Convolutional Networks (FCN) (Shelhamer et al. 2017), Deconvolution Network (Noh et al. 2015), and U-Net (Ronneberger et al. 2015). In this study, we developed a model based on U-Net, utilizing the encoder part of VGG16 (Simonyan and Zisserman 2015) with its fully connected layers removed (Fig. S1A).

The developed model was trained using supervised learning. To create the dataset for training, actual wheal images from urticaria patients were manually segmented under the supervision of four dermatologists. We labeled 31 training data samples. Before training, data augmentation (3000 images) was applied to expand the dataset(Fig. S1B). Fig. S1C shows that our model achieved a high level of accuracy sufficient to gain the approval of the supervising dermatologists.

### B Classification machine learning model of wheal types based on persistent images: PI-based Skin Eruption Classification Model (PI-SEclass Model)

The dataset used for machine learning of persistent images (PIs) was created using the following flow.

**1.** Six dermatologists independently classified the images of 105 patients into five types.
**2.** We selected the images classified with 4/6 reliability, namely, four dermatologists among six dermatologists agreed on the wheal type image into the same pattern.
**3.** Each selected image was labeled as A(Annular type), B(Broken-annular type), C(Circular type), D(Dot type), or G(Geographic type), and converted into grayscale images using
**4.** To minimize the influence of the imaging range (e.g., close-up shots of wheals or full-back images), five types of composed images were generated by size scaling the original images by 1x, 2x, 3x, 5x, and 7x, as shown in Fig. S2.
**5.** The grayscale images generated using the above method were augmented in the same manner as during segmentation, excluding brightness adjustment, to increase the number of images for the dataset.
**6.** Finally, we constructed a model that takes these grayscale images as input and outputs the classification of five types of wheals by modifying the final Dense layer of VGG-16 to have five-dimensional outputs. We trained the Dense layer of this model and confirmed that the training progressed successfully (see Fig. 4 in the main text).

**Figure S1:**
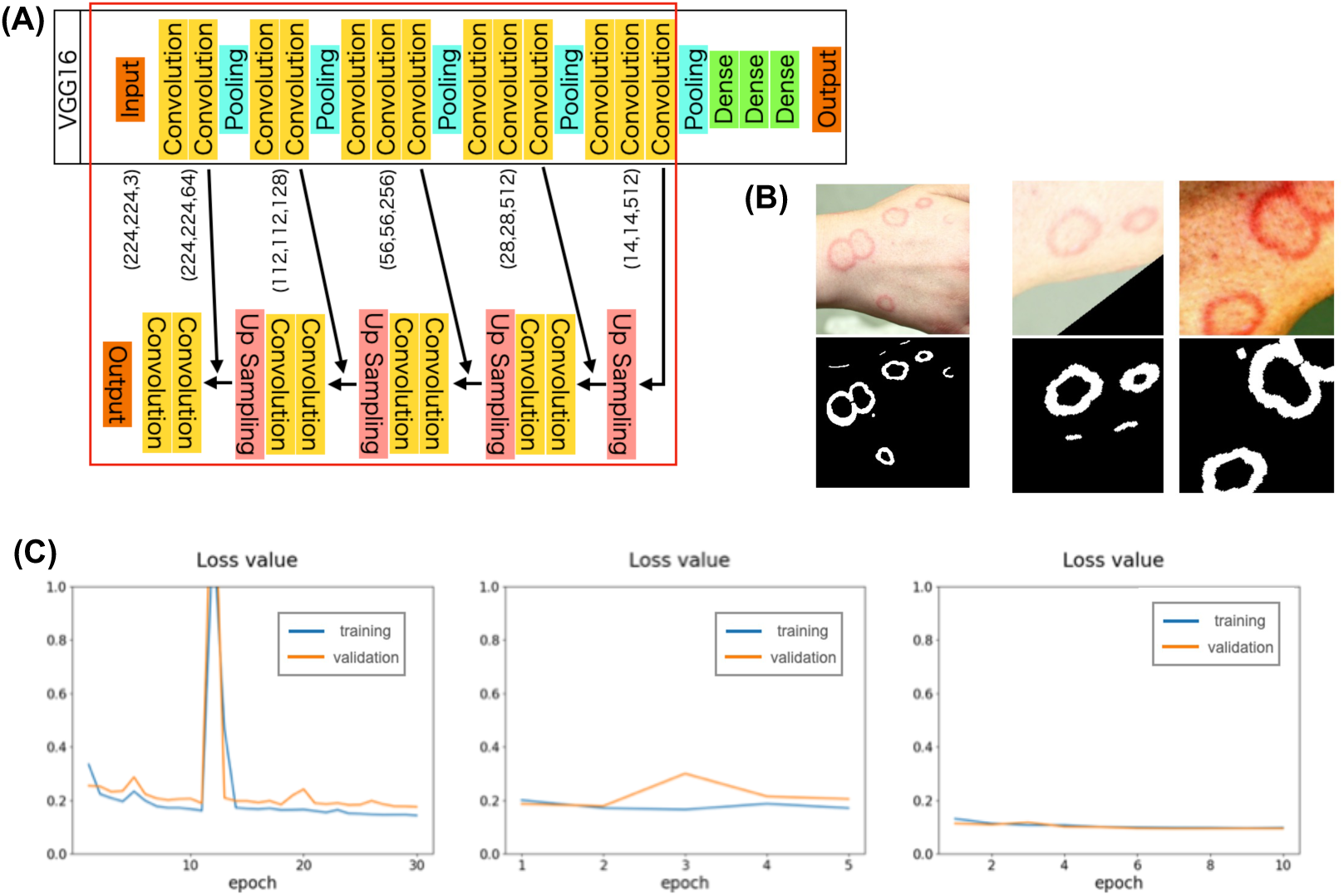
Segmentation model (SEseq model) overview. (A)The red frame indicates the model we developed. The output just before the fully connected layers of VGG16 is used as the input to the decoder layers. The number of filters in each decoder layer was set to match those in the VGG16 encoder: 512 for the (28,28) image size, followed by 256, 128, and 64. The activation function used is ReRU, and the loss function employed is Cross Entropy Loss. (B)The original patient image (upper panels) and the corresponding labeled training data image created (low panels). Two images generated from the image in the upper panels through Data Augmentation. Data Augmentation involves random rotation, scaling, gamma correction for brightness adjustment, and mirroring with a probability of 1/2. This process allows for the generation of a wide variety of images from a single original image. (C)Training Results of the Developed Segmentation Model. The model training was conducted in three stages: (Left panel) Transfer Learning for 30 epochs. (Middle panel) Fine Tuning for 5 epochs with a low learning rate. (Right panel)Identifying low-accuracy images and performing Transfer Learning again for 10 epochs. In all stages, the training progressed without noticeable overfitting.

**Figure S2:**
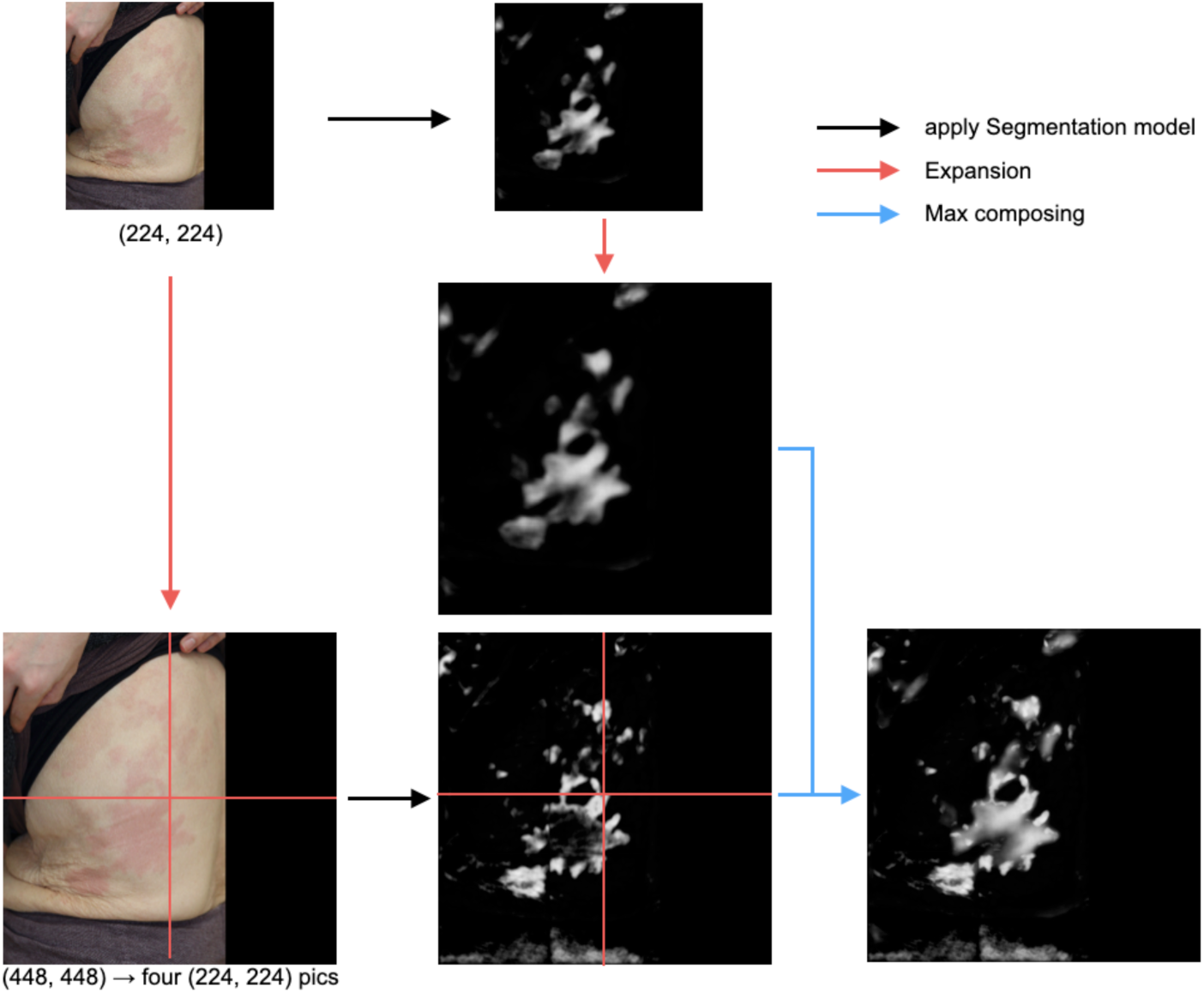
Example of Enlarged Composition (2x scale). The patient’s image was enlarged at the specified magnification, and both the original and enlarged images underwent segmentation. After resizing the images to match the same dimensions, they were combined using the maximum value method. In this study, images at magnifications of 1x, 2x, 3x, 5x, and 7x were all composed. Specifically, segmentation was performed for each magnification, the images were resized to the same dimensions, and the five resulting images were combined using the maximum value method. This approach enables effective handling of images captured over a wide range, whether they are close-up shots of wheals or broader views of the affected area. our segmentation model, SEseg-model.

### C Local sampling from global sampling

We first selected the parameter ranges globally. Since each wheal pattern has its own characteristic parameter range distinct from those of other patterns, we narrowed down the specific ranges for each pattern and parameter based on box plot analyses, as shown in Fig. S3. To generate more data for each pattern, we then selected parameter ranges corresponding to the values within the boxes.

### D The effect of wheal sizes on persistent images

To evaluate whether the selection of time points in *in silico* data influences the topological characteristics of wheal analysis using persistent homology, we performed a comparison of *in silico* wheal data obtained at two different time points through persistent homology analysis. Fig. S4 shows that the size effect of the wheal with the same parameter set appears in the features observed of persistent homology. Thus, we generate *in silico* repository for several time points with the same parameter sets.

**Figure S3:**
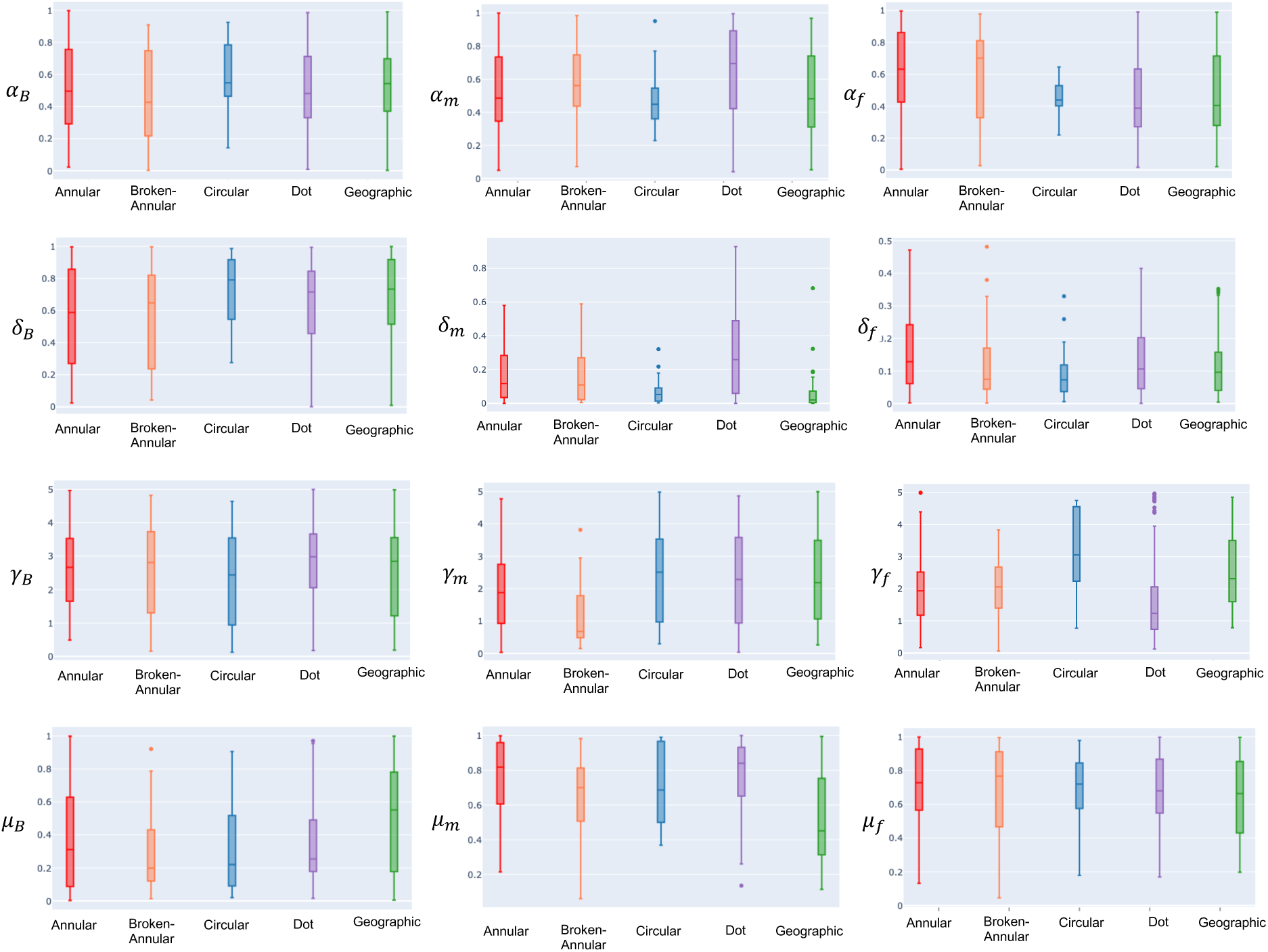
Box plots of parameter values obtained from global sampling. For local sampling, we select parameter ranges corresponding to the values within the boxes.

**Figure S4:**
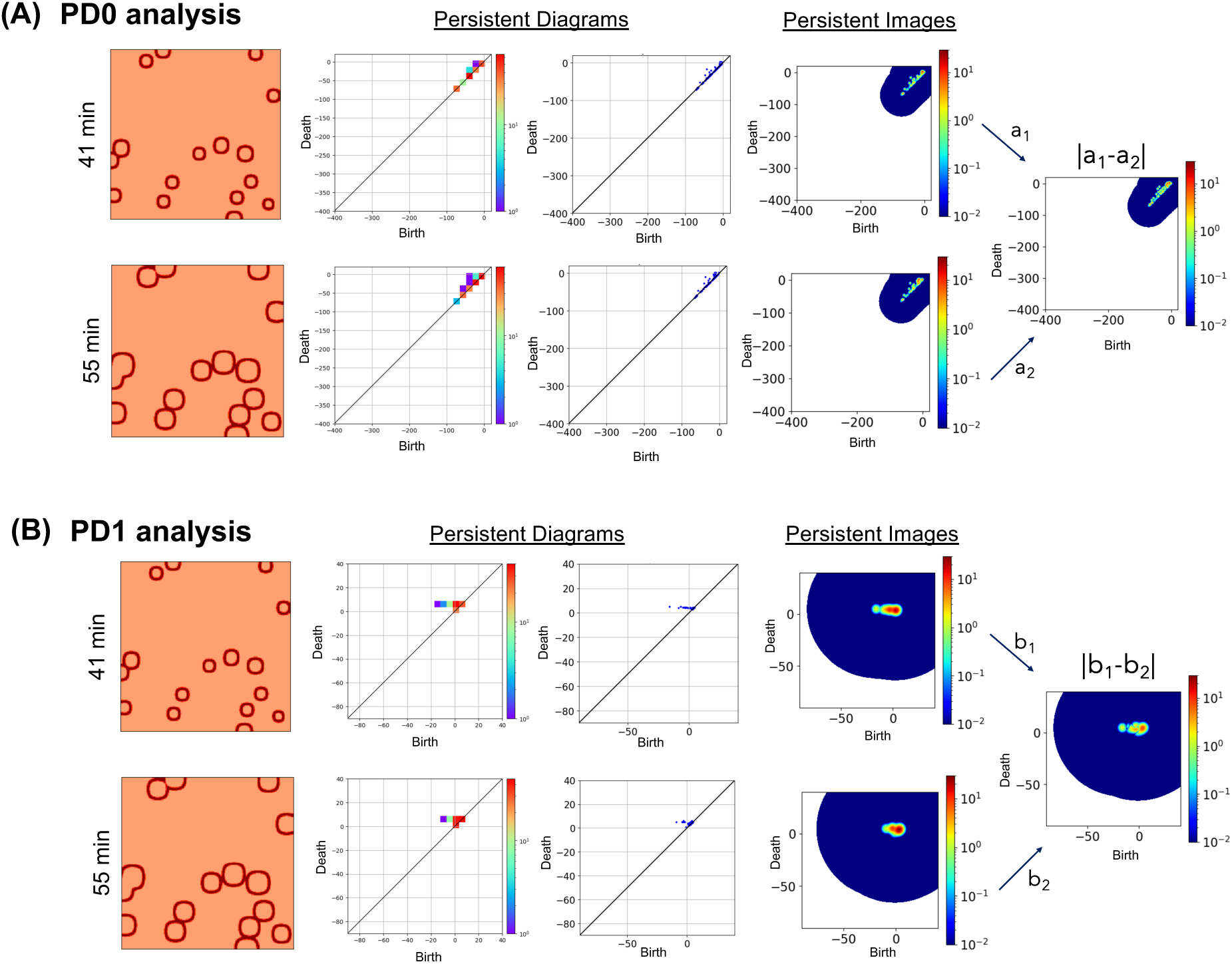
Simulation results between two different time points of *in silico* wheal data. (A-B) Results of persistence diagrams and images. The left panel of the persistence diagrams is plotted with frequency, while the right panel displays the persistence diagrams themselves. The final panel shows the absolute difference in persistence image values between the 41-minute and 55-minute data.

